# Evaluation of emotional arousal level and depression severity using the centripetal force derived from voice

**DOI:** 10.1101/2020.08.19.20177048

**Authors:** Shuji Shinohara, Hiroyuki Toda, Mitsuteru Nakamura, Yasuhiro Omiya, Masakazu Higuchi, Takeshi Takano, Taku Saito, Masaaki Tanichi, Shuken Boku, Shunji Mitsuyoshi, Mirai So, Aihide Yoshino, Shinich Tokuno

## Abstract

In this research, we propose a new voice feature called centripetal force (CF) to investigate the relationship between emotional arousal level and depression severity. First, CFs were calculated from various speech recordings in the interactive emotional dyadic motion capture database, and the correlation with the arousal level of each voice was examined. The resulting correlation coefficient was 0.52. We collected a total of 178 datasets comprising 10 speech phrases and the Hamilton Rating Scale for Depression (HAM-D) score of outpatients with major depression at the Ginza Taimei Clinic (GTC) and the National Defense Medical College (NDMC) Hospital. The correlation coefficients between CF and HAM-D scores were –0.33 and –0.43 at the GTC and NDMC, respectively. Next, the dataset was divided into the no depression group (HAM-D< 8) and the depression group (HAM-D ≥8) according to the HAM-D score. There was a significant difference in the mean CF values between the two groups in both the GTC and NDMC data (p = 0.0089 and p = 0.0016, respectively). The AUC when discriminating both groups by CF was 0.76 in GTC data and 0.72 in NDMC data. Indirectly, using CF established a relationship between emotional arousal level and depression severity.

## Introduction

The relationship between depression and emotional arousal has been established through studies using neurophysiological tests such as electroencephalogram (e.g., late positive potential amplitude), magnetoencephalography (e.g., neuromagnetic oscillatory activity), and skin conductance response [1–3]. Consequently, with the explosive increase in smartphone usage, research on voice emotion recognition and measurement of emotional arousal level using voice have been encouraged. For example, the relationship between arousal level and voice intensity or pitch has been documented [4,5].

The voice of a depressed person has dull, monotonous, and lifeless [6] features, and listeners can perceive patients’ distinctive prosody [7,8]. The Hamilton Rating Scale for Depression (HAM-D) [9] and self-administered questionnaires such as the General Health Questionnaire [10] and the Beck Depression Inventory [11] are powerful tools for measuring depression. However, if it becomes possible to measure depression severity by voice, through daily and remote monitoring, using the voice call feature of a smartphone. From this perspective, several studies have been conducted to measure the severity of depression using voice [12]; these have shown that speech characteristics are effective predictors of the signs and severity of depression [13].

Similarly, Cannizzaro et al. [14] examined the relationship between the HAM-D score and voice, and found a strong correlation between the HAM-D score and speaking rate or pitch variation. Yang et al. [8] demonstrated that changes in the severity of depression measured by the HAM-D, can be captured by the switching pause, that is, the pause duration between the end of one speaker’s utterance and the start of an utterance by the other. The mel-frequency cepstral coefficient (MFCC) is often used for voice recognition. Taguchi et al. [15] showed that MFCC2 (the second dimension of MFCC) is effective in classifying patients with depression and individuals without depression.

In a previous study, we found that the higher the arousal level, the higher were both the Hurst exponent and the zero-crossing rate of the waveform [16]. Specifically, we have shown that arousal level can be approximated by a weighted average of the Hurst exponent and zero-crossing rate. The zero-crossing rate is the rate at which the signal crosses the reference line, and is small in a smooth curve such as a sine curve and large in a rough waveform such as white noise. However, the Hurst exponent is expressed as 2-D, where D is the fractal dimension that represents the complexity of the waveform. In other words, the Hurst exponent is a measure of smoothness, which is the opposite of the fractal dimension, and theoretically it is 0 for white noise and 0.5 for brown noise.

In this paper, we propose a new speech feature called centripetal force (CF) that combines both the roughness and smoothness of the waveform. Next, CF was calculated from the emotional speech recordings stored in the interactive emotional dyadic motion capture (IEMOCAP) database [17], and compared with the arousal level evaluated by the annotators. Next, CF was calculated from the voice of the depressed patient and compared with the HAM-D score.

## Methods and Materials

### Acquisition of Data

#### Data on emotional arousal level

We used the IEMOCAP database [17] to investigate the relationship between the proposed voice index, CF, and emotional arousal. The database contains audio recordings of dyadic mixed-gender pairs of voice over artists. This database contains voices for five sessions in total, that is, five male voices and five female voices. The voices were manually divided into utterances (i.e., the sounds/words made from one breath to the next). There were 10,039 utterances in total. The arousal level of each utterance was evaluated by at least two different annotators on a five-point scale. The arousal level of each utterance was calculated as the average of the evaluation values given by each annotator.

Furthermore, emotion categories were evaluated by at least three annotators. There were nine emotion categories: “angry,” “happy,” “sad,” “neutral,” “frustrated,” “excited,” “fearful,” “surprised,” and “disgusted.” However, utterances that did not seem to fit into any of these categories were classified as “other.” A simple majority voting method was used to assign an emotion category if there was disagreement among annotators regarding classification. The annotators were also allowed to tag more than one emotion category. If no majority category could be assigned, the category was labeled xxx. The total number of utterances assigned to the nine emotion categories by the above procedure was 7,527.

#### Data on severity of depression

This study collected data from outpatients with major depressive disorder after obtaining written informed consent from all participants at the Ginza Taimei Clinic (GTC) and the National Defense Medical College (NDMC) Hospital. At each health care facility, the recruited patients were instructed to pronounce 17 Japanese phrases. However, the 17 phrases collected at the two hospitals were not exactly the same. Of the 17 phrases, 10 were common. For the purpose of this study, these 10 phrases were used for analysis. Table 1 shows the contents of these 10 phrases.

**Table 1.** Ten phrases used for analysis

| **Phrase**<br>**number** | **Phrase in Japanese** | **Purpose (meaning)** |
| --- | --- | --- |
| P1 | I-ro-ha-ni-ho-he-to | Non-emotional (similar to “a-b-c”) |
| P2 | Honjitsu ha seiten nari | Non-emotional (It is fine today) |
| P5 | Mukashi aru tokoro ni | Non-emotional (Once upon a time, there lived) |
| P11 | Garapagosu shotou | Check pronunciation (Galápagos Islands) |
| P12 | Tsukarete guttari shiteimasu. | Emotional (I am tired/dead tired) |
| P13 | Totemo genki desu | Emotional (I am very cheerful) |
| P14 | Kinou ha yoku nemuremashita | Emotional (I was able to sleep well yesterday) |
| P15 | Shokuyoku ga arimasu | Emotional (I have an appetite) |
| P16 | Okorippoi desu | Emotional (I am irritable) |
| P17 | Kokoroga odayaka desu | Emotional (My heart is calm) |

Voice was recorded using a pin microphone (ME52W, Olympus, Tokyo, Japan) attached to the patient’s chest, approximately 15 cm from the mouth. The recording equipment was a portable recorder R-26 (Roland, Shizuoka, Japan). The record format involved a linear pulse-code modulation (PCM). The sampling frequency and number of quantization bits were 11,025 Hz and 16, respectively.

We also had a doctor interview each patient and provide a score on the HAM-D, in the same session the voice recordings were used, to evaluate the severity of depression in patients. In this way, pairs of recorded voices of 10 phrases and HAM-D scores were collected from 178 patients. Table 2 shows the participants’ information from each health care facility. Patients were included if they had been diagnosed with major depressive disorder (MDD) according to the Diagnostic and Statistical Manual of Mental Disorders, Fourth Edition, Text Revision [18] and were aged over 20 years. They were excluded if they had been diagnosed with serious physical disorders or organic brain disease. They were diagnosed by a psychiatrist using the Mini-International Neuropsychiatric Interview [19].

**Table 2.** Participants’ information

| Health<br>care<br>Facility | Sex | Number of<br>participants | | Mean age ± SD | |
| --- | --- | --- | --- | --- | --- |
| GTC | Female | 55 | 88 | 31.6 ± 8.6 | 32.0 ± 7.9 |
| | Male | 33 | | 32.5 ± 6.5 | |
| NDMC | Female | 44 | 90 | 62.0 ± 13.1 | 55.2 ± 14.8 |
| | Male | 46 | | 48.8 ± 13.5 | |

The protocol of this study was designed in accordance with the Declaration of Helsinki and relevant domestic guidelines issued by the concerned authorities, in Japan. The protocol was approved by the ethics committee of the Faculty of Medicine, the University of Tokyo (no. 11572), and the ethics committee of the National Defense Medical College (no. 2248).

#### Proposed Method

Consider a signal *x(t),0≤ t #x2264; T*. The velocity *v(t)* of the signal at time “t” is defined as follows:

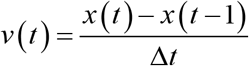

Next, the force *F(t)* that acts virtually is defined as follows:

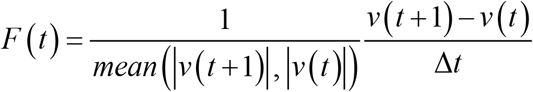

where *mean(x,y)*represents the mean of x and y and is used for normalization. In this study, the harmonic mean is used as the mean. For simplicity, we set *Δt* = 1. At this time, equation (2) can be rewritten as follows:

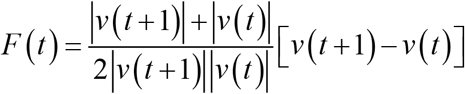

Next, we focus on the directionality of *F(t)*. The center line is defined as the average time 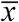 of the signal as follows:

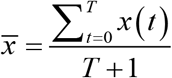

In the case of *F*(t)<0, F(t) represents the downward force. Therefore, when *x(t)* is above the center line, i.e., when 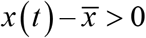 *F(t)* represents the force toward the center. Correspondingly, in the case of 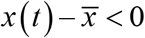 *F(t)* becomes a force away from the center. In addition, in the case of *F(t)*>0, *F(t)* represents an upward force; when 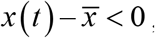 *F(t)* represents the force toward the center. Accordingly, when 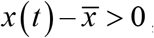 *F(t)* represents the force away from the center. Centripetal force (CF) is the force which toward the center is positive and away from the center is negative (Figure 1a). It is defined as follows:

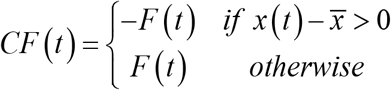

**Figure 1a.**
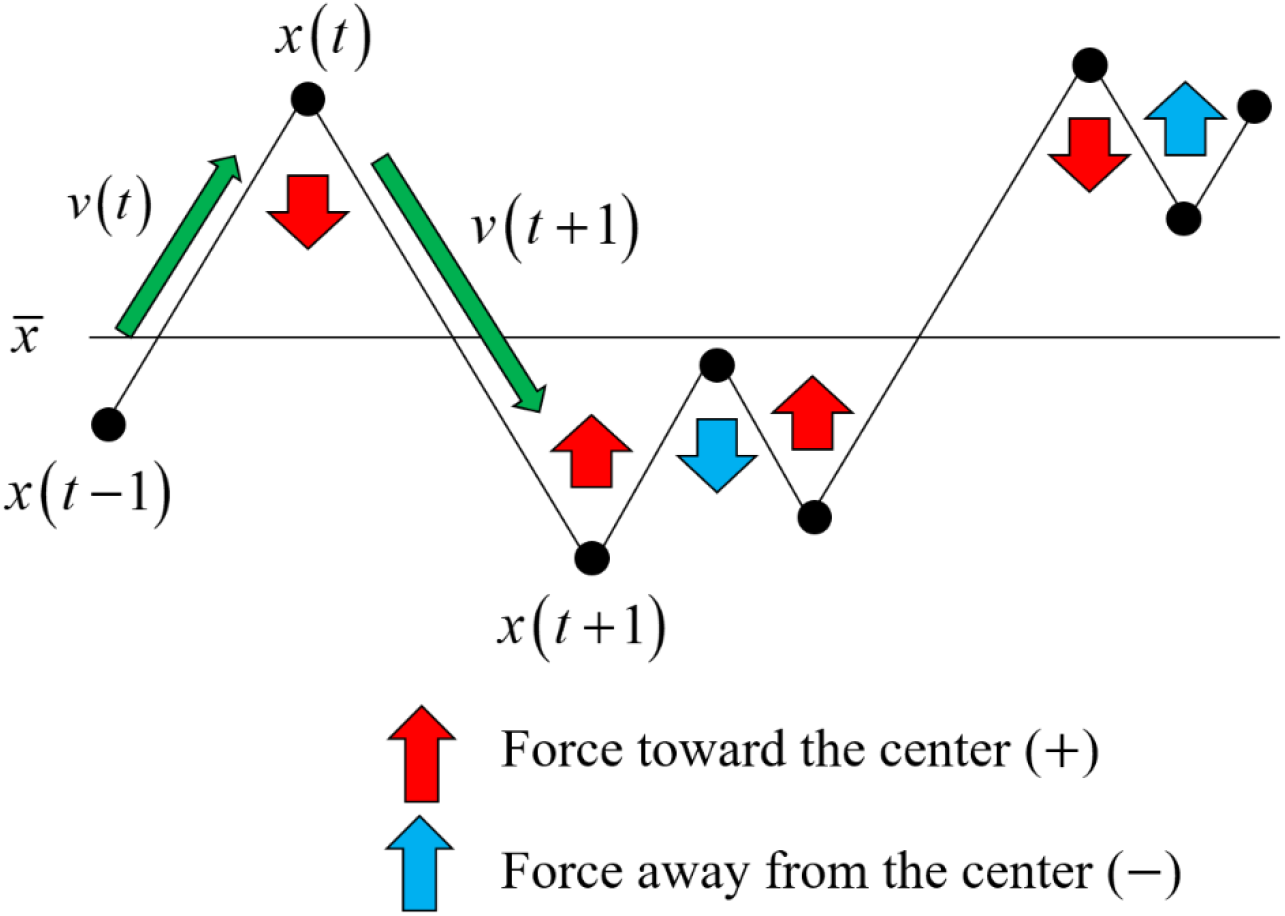
Direction of centripetal force.

*Note*The force toward the center is positive, and the force away from the center is negative.

The CF of a given voice signal is defined as the average time of *CF(t)* as follows:

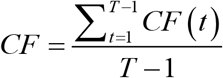

As is clear from the definition, CF increases as the proportion of force toward the center increases. In addition, as can be seen by comparing Figures 1a and 1b, the higher the ratio of the waveform crossing the center line, that is, the higher the zero crossing ratio, the larger the ratio of the force toward the center. Thus, CF can be said to be a measure that reflects the roughness of the waveform, like the zero-crossing rate.

Next, we focus on the first term 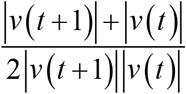 on the right side of the equation (3). This term is the reciprocal of the harmonic mean of *|v(t)|* and *|v(t+1)|* The harmonic mean of x and y is expressed as 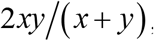, and the more skewed the values of x and y, the smaller the harmonic mean. Conversely, the more biased the values of *|v(t)|* and *|v(t+1)|*, the greater the CF, and this increases as the bias between them increases. As can be seen by comparing Figures 1b and 1c, the larger the deviation between the values of *|v(t)|* and *|v(t+1)|*, the smoother the entire waveform tends to be. In other words, the first term on the right side of equation (3) can be considered as a measure of smoothness. Thus, CF can be regarded as a measure that reflects roughness and smoothness.

**Figure 1b.**
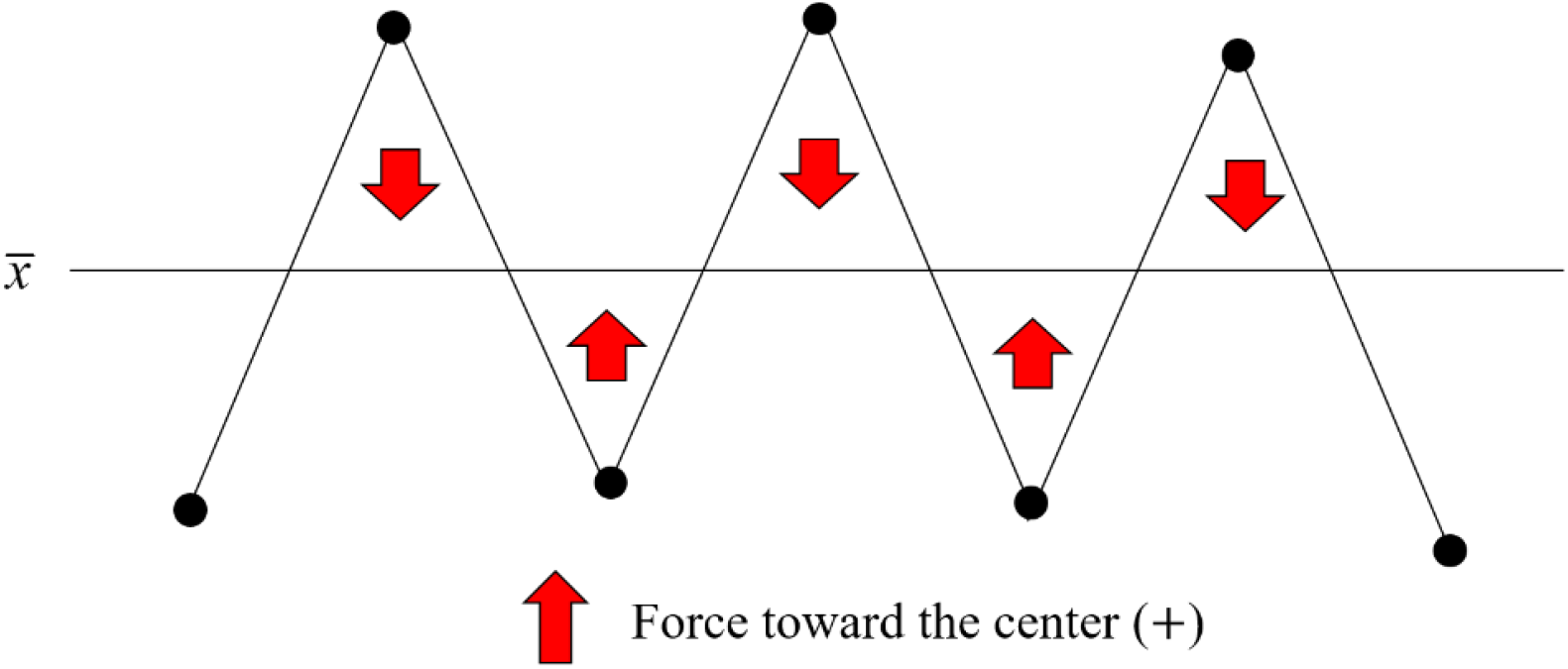
Centripetal force for rough waveform.

*Note:* The more the waveform crosses the centerline, the greater the proportion of force towards the center.

**Figure 1c.**
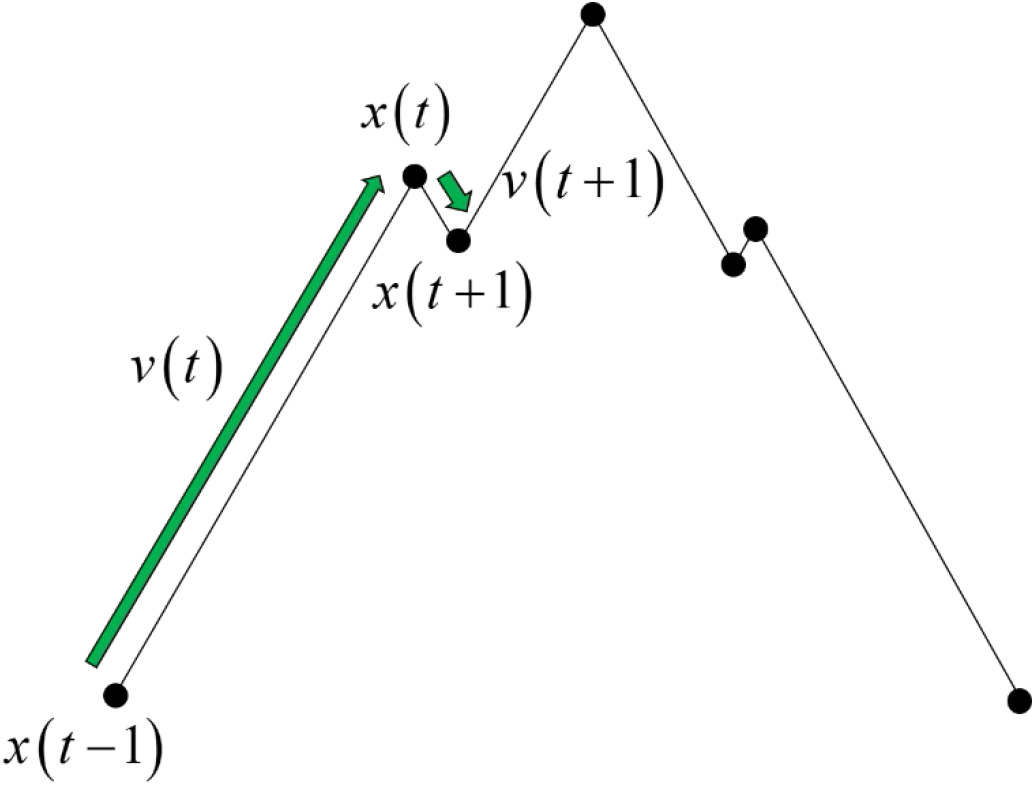
Waveform when there is a bias between two adjacent velocity vectors.

*Note*When the absolute values *|v(t)*| and *|v(t+1)|* of two adjacent velocity vectors differ greatly, the entire waveform tends to be smooth.

## Results

### CF as an arousal level index

CFs were calculated from 10,039 utterances stored in the IEMOCAP database. The correlation coefficient between CF and the arousal level of each utterance given by the annotators was 0.52 (n = 10039, p < 2.2×10^−16^). We extracted low arousal voices with a level of 2 or less (n = 1112, mean ± SD = 1.92 ± 0.19) and high arousal voices with a level of 4 or more (n = 1692, mean ± SD = 4.19 ± 0.28), from the database. As a result of distinguishing these two groups by CF, the area under the curve (AUC) of the receiver operating characteristic curve was 0.93 and when the cutoff value was 0.90, both the sensitivity and specificity were 0.86.

The average values of arousal level and CF for each emotion category were compared. There were nine emotion categories: “angry,” “happy,” “sad,” “neutral,” “frustrated,” “excited,” “fearful,” “surprised,” and “disgusted.” Figure 2 shows the number of utterances in each category. In addition, Figure 3 shows the average values of arousal level and CF for each category. Here, the categories on the horizontal axis are arranged in descending order of average arousal level. Except for the three categories of “fearful,” “surprised,” and “disgusted,” the order relationship between arousal level and CF was the same. As can be seen from the figure, both arousal levels and CF tended to be high in the categories of “angry” and “excited,” and low in the categories of “neutral” and “sad,” which is consistent with our daily feelings. The number of utterances included in the three categories, in which the order of arousal level and CF did not match, was extremely small, compared to other categories. The utterances (percentage) of the categories “fearful,” “surprised,” and “disgusted” were 40 (0.53%), 103 (1.42%), and 2 (0.03%), respectively. The above analysis was performed using the statistical software R [20]. All analyses were performed using the statistical software R, unless otherwise specified.

**Figure 2.**
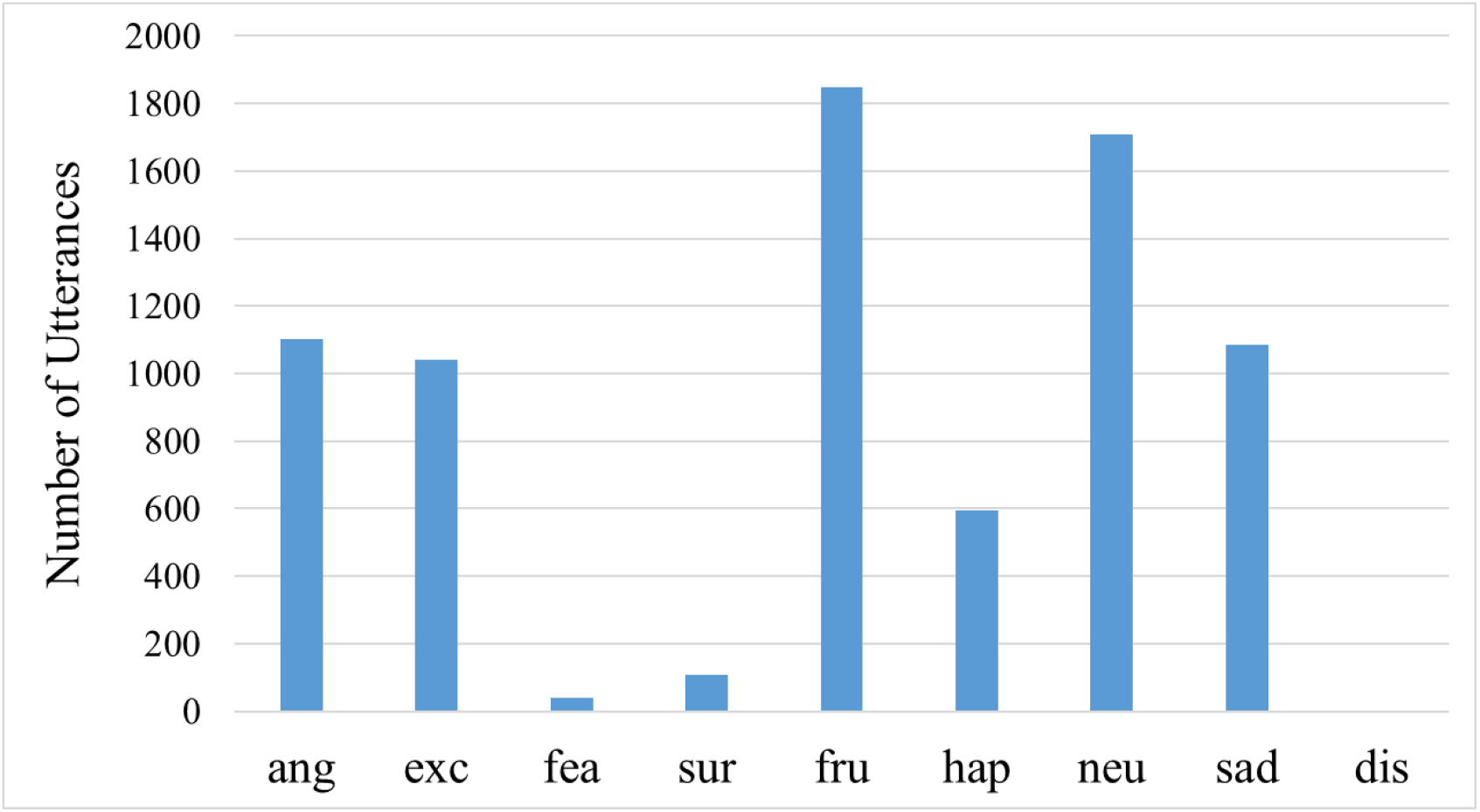
The number of utterances for each emotion category.

*Note:*The number of utterances classified in “disgusted” was 2. Additionally, ang = angry; exc = excited; fea = fearful; sur = surprised; fru = frustrated; hap = happy; neu = neutral; sad = sad; dis = disgusted.

**Figure 3.**
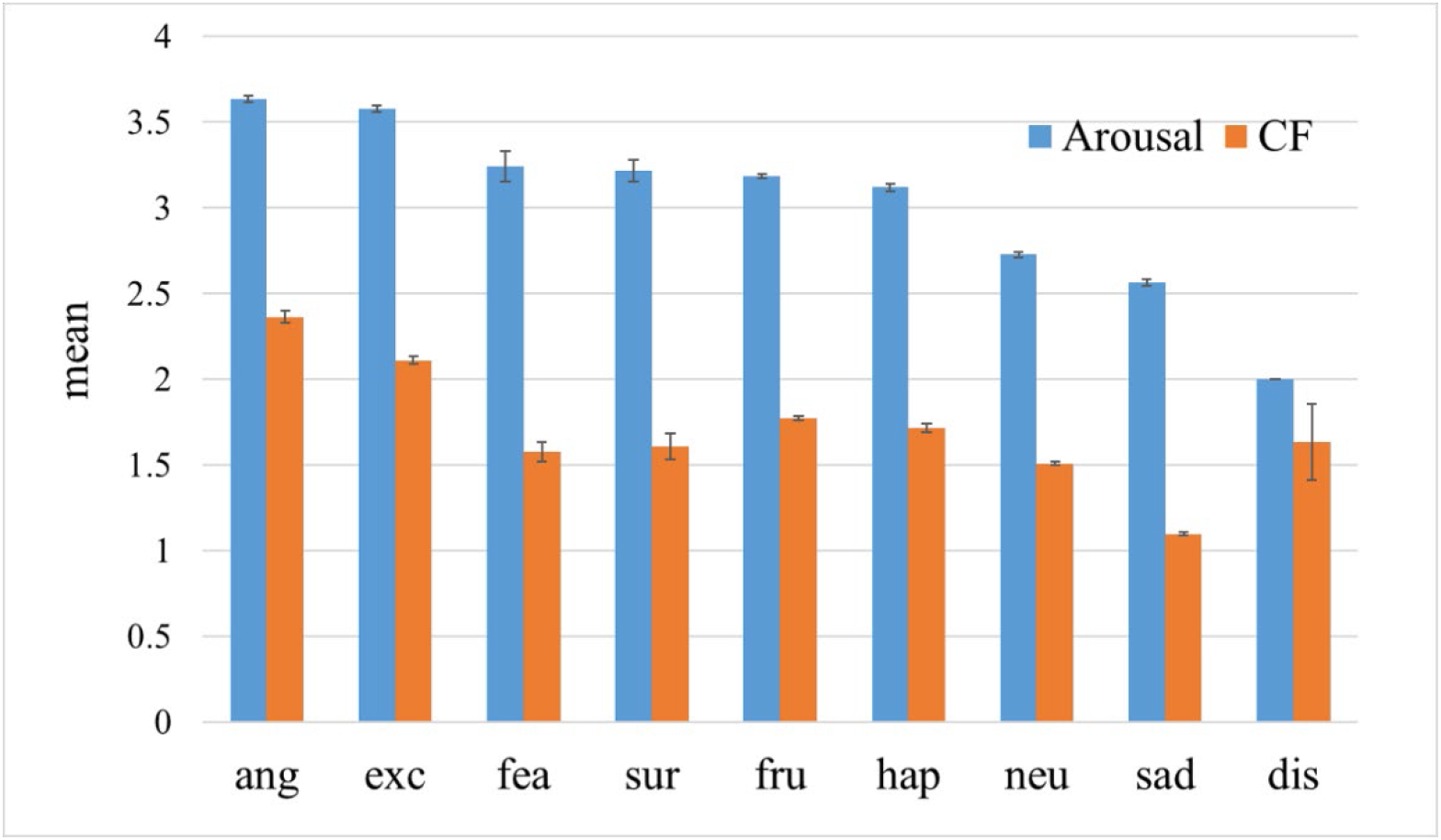
The mean arousal level and the mean CF for each emotion category.

*Note:*Error bars represent standard error. Additionally, ang = angry; exc = excited; fea = fearful; sur = surprised; fru = frustrated; hap = happy; neu = neutral; sad = sad; dis = disgusted.

### HAM-D score

There are various discussions on the classification of depression severity using the HAM-D score [21]. In this study, the dataset was divided into two groups, a “no depression” group with a HAM-D score of less than 8 and a “depression” group with a HAM-D score of 8 or more, using Hashim’s method [22]. Table 3 shows the mean HAM-D score of each group according to the facility. According to the Wilcoxon rank sum test, a significant difference was found between the HAM-D scores of each group, in both the GTC and the NDMC Hospital (p = 1.22×10^−8^, p = 1.56×10^−13^, respectively).

**Table 3.**
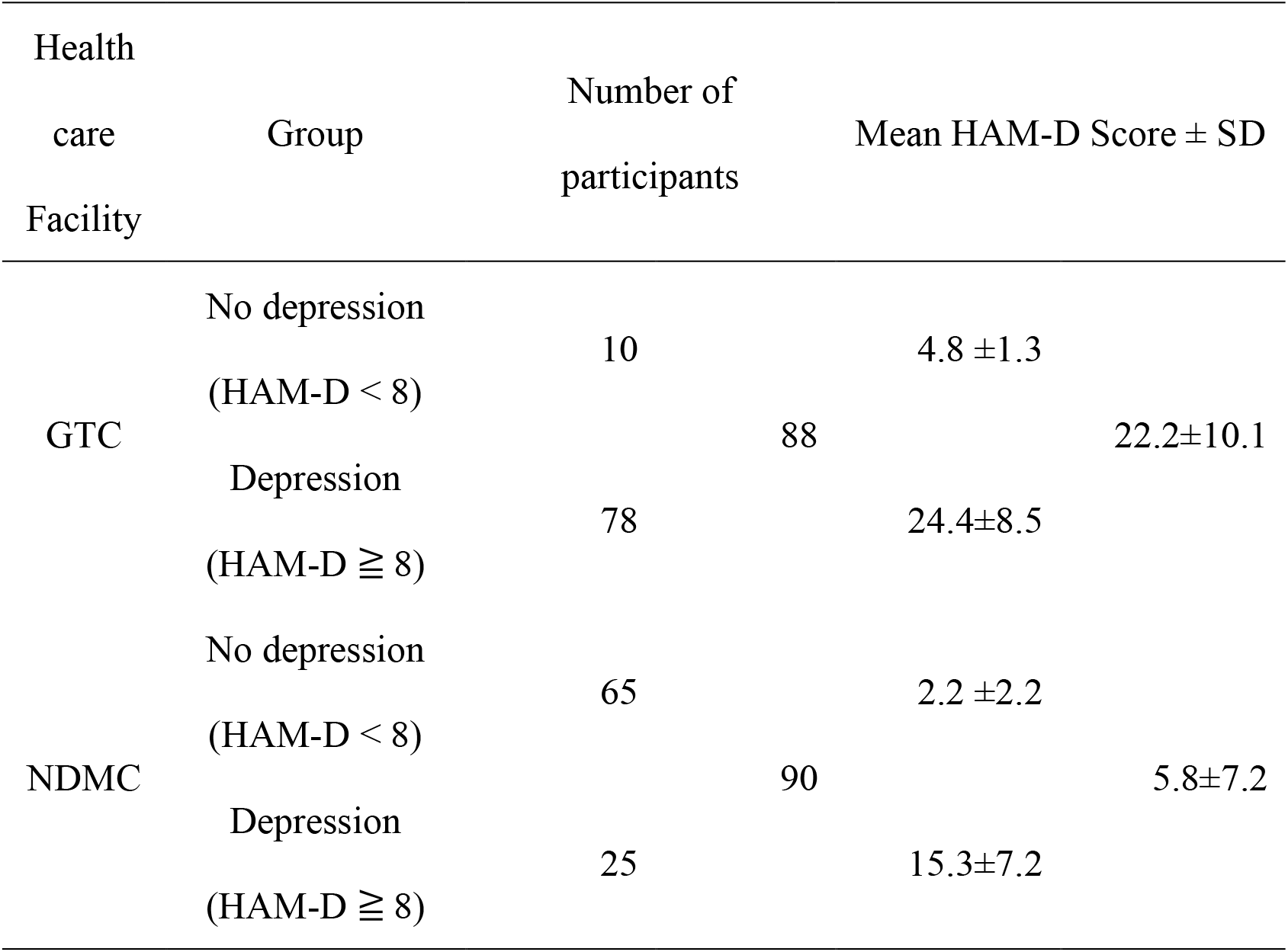
Mean scores on the Hamilton Rating Scale for Depression (HAM-D)

### CF as an index of depression severity

Data of patients with major depression were collected from two health care facilities. As shown in Table 2, the age groups of patients at both facilities were quite different. In addition, the sound field environments may be different in both facilities. Therefore, a separate analysis was performed for each hospital. Figure 4 shows the mean CF values of each group for each facility. However, the CF value of each participant was obtained by calculating the mean value of the CFs for the 10 phrases. At the GTC, the mean CF values of the “no depression” group and “depression” group were 0.67 ± 0.041 (n = 10) and 0.54 ± 0.015 (n = 78), respectively. At the NDMC Hospital, the mean CF values of the “no depression” and “depression” groups were 0.81±0.020(n = 65) and 0.69±0.031 (n = 25), respectively.

**Figure 4.**
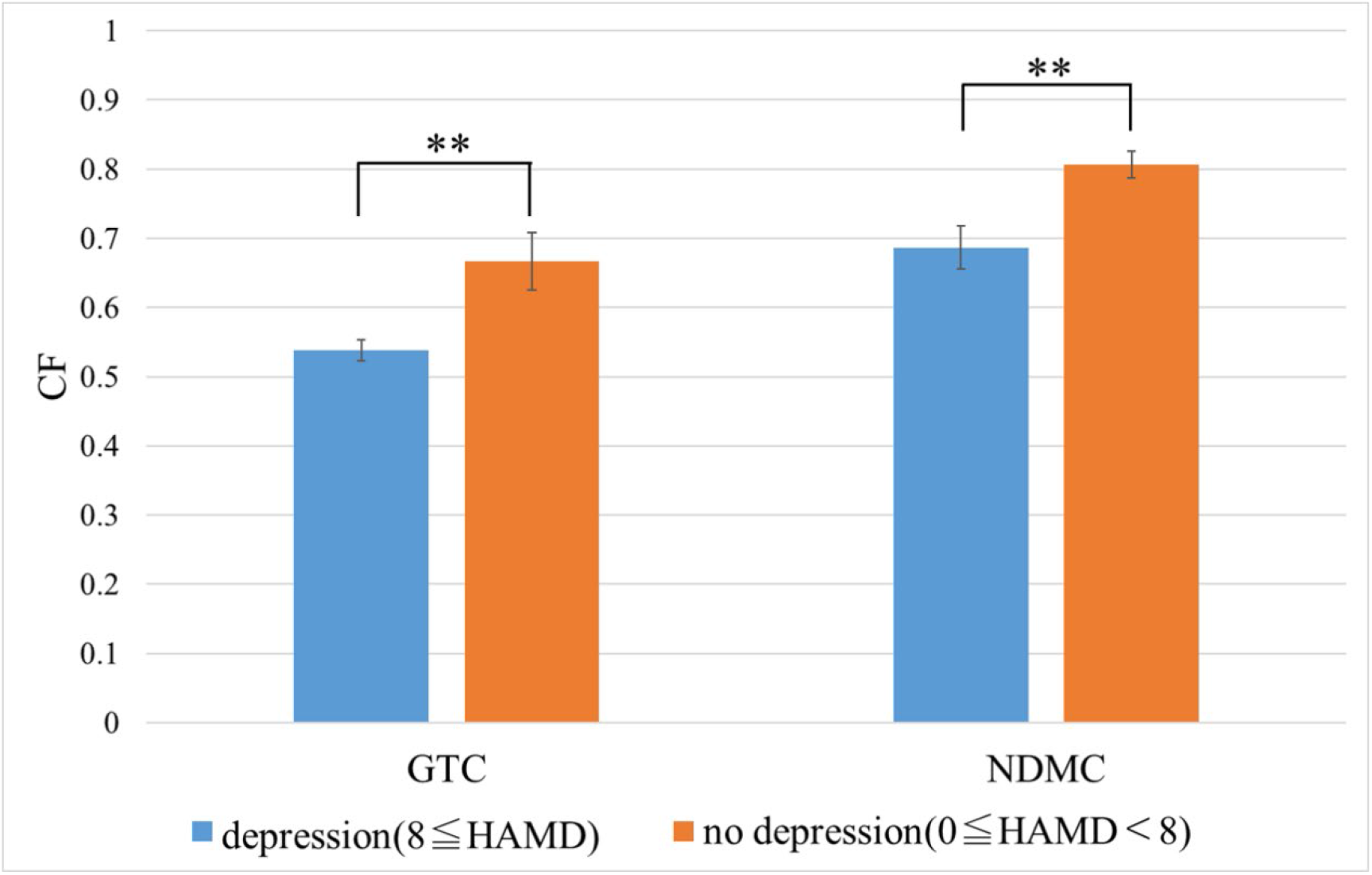
The mean CF for the “depression” and “no depression” groups, by facility.

*Note:*Error bars represent standard error. ** (p < 0.01)

As a result of the Wilcoxon rank sum test, a significant difference was found between the mean CF values of each group in both the GTC and the NDMC Hospital (p =0.0089 and p = 0.0016, respectively). AUC when the groups were identified using CF was 0.76 for the GTC (cutoff point = 0.62, sensitivity = 0.80, specificity = 0.73) and 0.72 for the NDMC Hospital (cutoff point = 0.63, sensitivity = 0.89, specificity = 0.48). The correlation coefficient between the HAM-D score and the CF was –0.33 (n = 88, p = 0.0018) in the GTC and –0.43 (n = 90, p = 2.16×10^−5^) in the NDMC Hospital.

Next, we examined the effect of different phrases on the CF values obtained from each facility. A two-way analysis of variance (ANOVA) of CF was conducted for two factors: group (“no depression” and “depression”) and phrase (10 phrases).

At the GTC, there were significant differences in both group and phrase factors (F(1, 860) = 39.13, p = 6.24×10^−10^, F(9, 860) = 31.20, p< 2.00×10^−16^). However, there was no interaction between group and phrase (F(9, 860) = 0.32, p = 0.97).

At the NDMC Hospital, there were significant differences in both group and phrase factors (F(1,880) = 74.24, p< 2.00×10^−16^, F(9,860) = 16.11, p< 2.00×10^−16^). However, there was no interaction between group and phrase (F(9,860) = 0.15, p = 1.00). Figures 5a and 5b. show the mean CF for each phrase in the “no depression” and “depression” groups. In all phrases, CF values of the “no depression” group were higher than those of the “depression group” at both facilities.

**Figure 5a.**
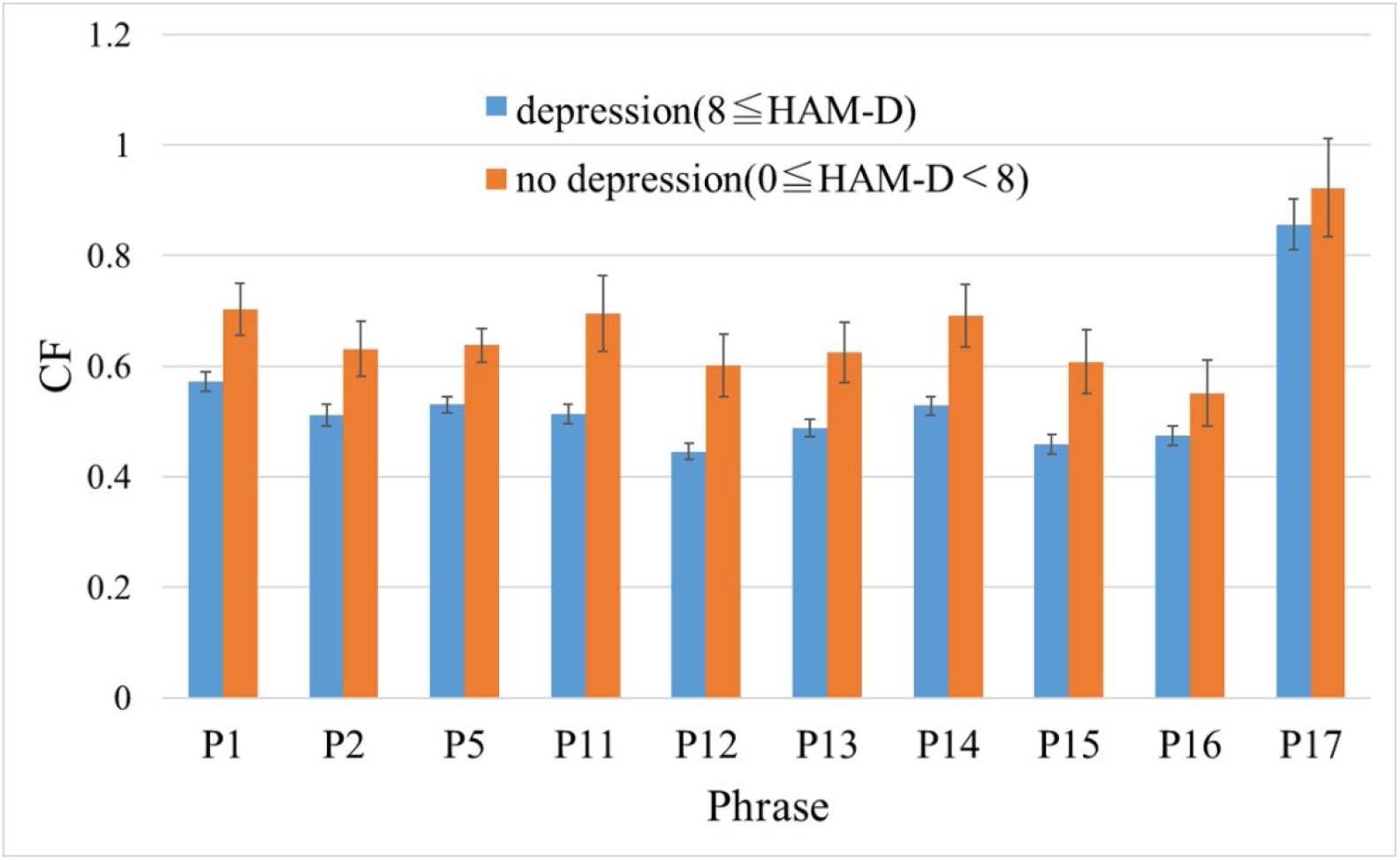
The mean of the CF values of the “no depression” and “depression” groups for each phrase at the GTC.

*Note:*Error bars represent standard error.

**Figure 5b.**
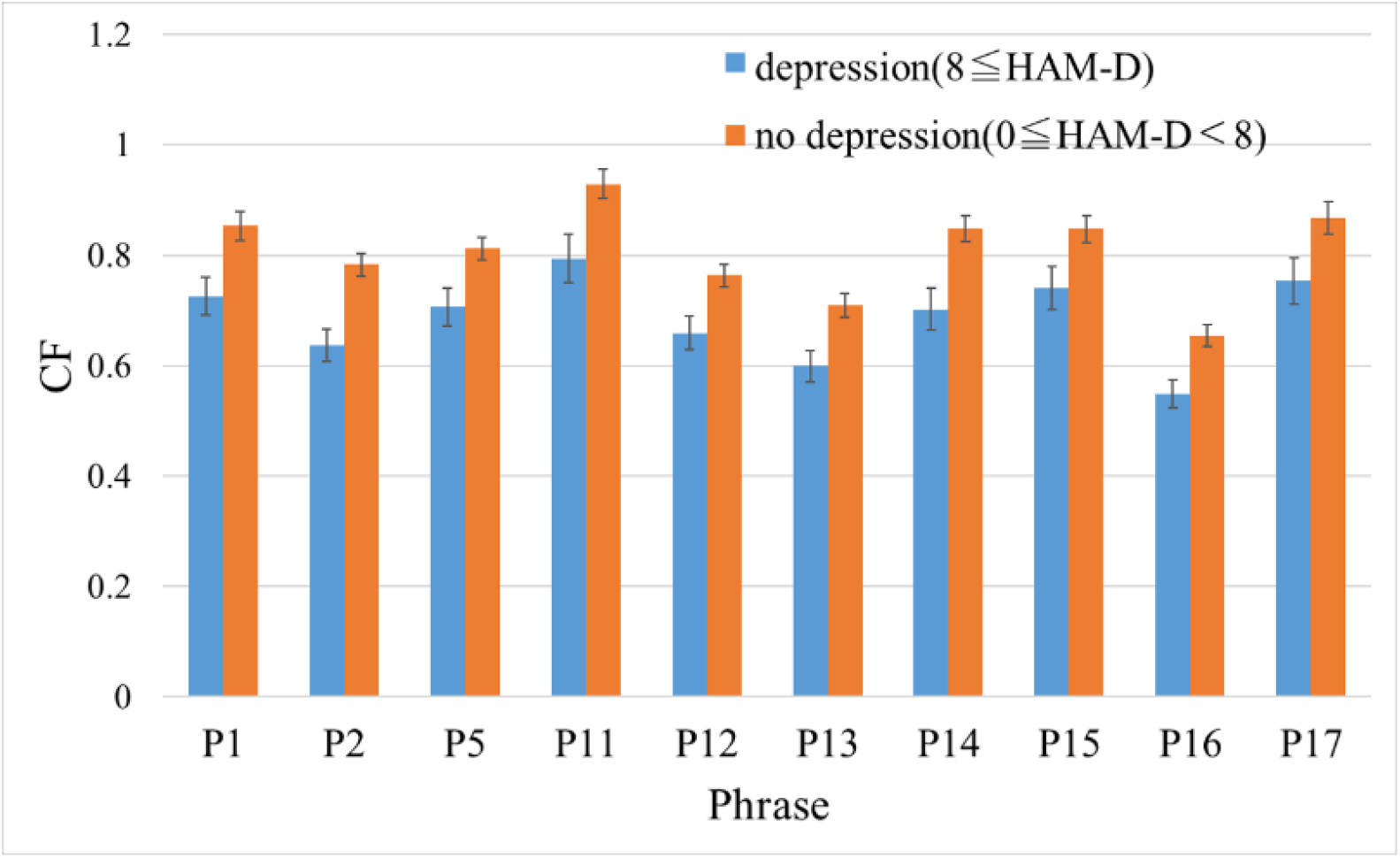
The mean of the CF values of the “no depression” and “depression” groups for each phrase at the GTC.

*Note:*Error bars represent standard error.

Table 4 shows a summary of classification performance between the “no depression” and “depression” groups by CF. The table shows the *p*-values obtained by the Wilcoxon rank sum test, the AUC, and the correlation coefficient between HAM-D and CF for each phrase of both facilities. In the table, the minimum *p*-value, the maximum AUC, and the maximum correlation for each hospital are shown in bold fonts. The AUC tended to be higher for the GTC. On the other hand, the correlation coefficient with HAM-D tended to be higher for the NDMC Hospital.

**Table 4.** A summary of classification performance between no depression and depression groups by CF

| Phrase | p-value<sup>a</sup> |  | Area Under the Curve (AUC) |  | Correlation coefficient |  |
| --- | --- | --- | --- | --- | --- | --- |
|  | GTC | NDMC | GTC | NDMC | GTC | NDMC |
| P1 | 0.019* | 0.0056** | 0.73 | 0.69 | -0.27* | -0.34** |
| P2 | 0.026* | **0.00015\*\*\*** | 0.72 | **0.76** | -0.28** | **-0.49\*\*\*** |
| P5 | **0.0073\*\*** | 0.0089** | **0.76** | 0.68 | -0.22* | -0.38*** |
| P11 | 0.0086** | 0.0053** | 0.76 | 0.69 | -0.25* | -0.37*** |
| P12 | 0.012* | 0.0058** | 0.75 | 0.69 | -0.25* | -0.34** |
| P13 | 0.017* | 0.0066** | 0.73 | 0.69 | -0.30** | -0.38*** |
| P14 | 0.0076** | 0.0019** | 0.76 | 0.71 | **-0.31\*\*** | -0.41*** |
| P15 | 0.019* | 0.016* | 0.73 | 0.67 | -0.26* | -0.35*** |
| P16 | 0.25 | 0.0024** | 0.61 | 0.71 | -0.21 | -0.39*** |
| P17 | 0.36 | 0.050* | 0.59 | 0.63 | -0.21* | -0.35*** |
| Total | 0.0089** | 0.0016** | 0.76 | 0.72 | -0.33** | -0.43*** |

\*\*\* (p < 0.001), \*\* (p < 0.01), \* (p < 0.05). <sup>a</sup> by Wilcoxon rank sum test.

## Discussion

The CF, a voice index proposed by this paper, showed a significant correlation with the arousal level evaluated by annotators. In the comparison of CF and arousal level by emotion category, the order relation between “fearful,” “surprised,” “frustrated,” and “happy” was reversed. However, as can be seen from Figure 3, the values of the arousal level are almost the same for these four emotions, and it can be said that it is difficult even for human beings to distinguish among them.

Regarding the emotion category “disgusted,” the values of the arousal level and CF were significantly different, but this may be because the sample size was too small (n = 2). With regard to depression severity, there was also a significant correlation between the HAM-D score and the CF value. The AUC was over 0.7 in both facilities. MFCC2 showed a very high AUC of 0.88 in discriminating healthy individuals from patients suffering from MDD [15]. However, there was no significant correlation between the MFCC2 and the severity of depression. On the other hand, the CF value significantly correlated with the HAM-D score. It should be noted that the AUC of CF was lower than that of MFCC2, but in this study, all the participants were outpatients suffering from depression. In future studies, we would like to compare the results of healthy individuals with those of patients suffering from depression.

The model classifying a depressive state versus a euthymic state had an AUC of 0.78 in the study by Faurholt-Jepsen et al. [23]. Here, a depressive state is defined as one with a HAM-D score ≥13 and Young Mania Rating Scale (YMRS) score < 13, while a euthymic state is defined as one with a HAM-D score < 13 and YMRS score < 13. Although no simple comparison is possible, the AUC shown here was about the same value as that of the CF. However, it may be advantageous because the CF is unlikely to be overfitted as it consists of only one feature.

As shown above, the CF was shown to be associated with arousal and depression severity. In other words, though indirectly, the relationship between arousal and depression severity was associated by the CF. However, it should be noted that the arousal level was an evaluation value given by the annotator and did not reflect the participant’s own evaluation. In the future, it is necessary to investigate the relationship between CF and physiological indicators.

In addition, we need to clarify the qualitative meaning of CF. The CF correlated with the arousal level, and was high for the voices expressing anger and excitement and low for those expressing sadness and neutral qualities. It is necessary to consider why a voice with a high arousal level increases the force toward the center.

This study has some limitations. First, all participants had to use the same phrases for an accurate evaluation. Our future task would be to analyze the CF of spontaneous speech. The two-way ANOVA revealed that CF was affected by the utterance content. Future studies should explore the reasons for such differences among phrases. Second, the sample size of the group was small. We collected voices from two health care facilities, the GTC and the NDMC Hospital, but the age group and the distribution of HAM-D scores were quite different at both facilities. The mean age of the participants was higher in the NDMC Hospital. Conversely, the HAM-D score was higher in the GTC. The reason for this may be that the GTC is located in central Tokyo and many outpatients are young office workers who commute to the city center; while the NDMC Hospital is in the suburbs and several patients are elderly people living in the locality. The NDMC Hospital is a university hospital and many patients have already been treated at other hospitals. Future studies should include a larger sample size acquired in the similar environments.

We developed a smartphone application for monitoring mental health using voice data from smartphone calls, called the Mind Monitoring System (MIMOSYS) [24], and conducted a large-scale verification experiment [25,26]. The results of these studies suggested that MIMOSYS may be effective for monitoring changes in mental health due to stress. Mental health monitoring is most effectively accomplished by performing long-term time-series analysis of the acquired data, considering the user’s life events. In the future, we plan to implement the CF algorithm in MIMOSYS and perform large-scale verification.

## Data Availability

According to Japanese law, the sensitivity of audio files is similar to that of any other personal information and cannot be published without consent. In this research protocol, we did not obtain consent from the participants to publish the raw audio files as a corpus. The datasets used and/or analyzed during the current study are available from the corresponding author upon reasonable request.

## Acknowledgements

This research was partially supported by the Center of Innovation Program from the Japan Science and Technology Agency (JST). This work was also supported by the JSPS KAKENHI, Grant Number JP16K01408. We would like to thank Editage (https://www.editage.com) for English language editing.

## Author Contributions

S. T. was responsible for the design of the clinical study. A. Y., H. T., T. S., and M. T. were responsible for the execution of the clinical study including patient recruitment, retention, and data collection. S. S. conceived the CF algorithm, analyzed the data, and wrote the manuscript. M. N., Y. O., M. H., T. T., S. B., S. M., and S. T. contributed to the interpretation of the study findings. All authors participated in the editing and revision of the final version of the manuscript.

